# Comparative Study on Image Quality of Deep Learning and Adaptive Statistical Iterative Reconstruction-V in Thin Layer CT of liver Lesions

**DOI:** 10.64898/2026.05.23.26353923

**Authors:** Jie Yang, Li Li, Jian Cao, Jing Zhang

## Abstract

**Objective:** This study aims to compare the advantages and disadvantages of DLIR and adaptive statistical iterative reconstruction-V (ASIR-V) in thin-slice (2.5 mm) CT images of hepatic lesions characterized by high and low contrast. Additionally, the study seeks to determine the optimal DLIR strength for the evaluation of liver lesions.

**Methods:** A retrospective analysis was performed on 90 patients who underwent abdominal contrast-enhanced CT scans. Group A comprised 48 patients with low-contrast lesions, while Group B included 42 patients with high-contrast lesions. The acquired images were reconstructed using post-processing DLIR at low (DLIR-L), medium (DLIR-M), and high (DLIR-H) strengths, all with a slice thickness of 2.5 mm (subgroups A1-A3, B1-B3). Furthermore, images were reconstructed with ASIR-V at 50% strength at slice thicknesses of 2.5 mm and 5 mm (subgroups A4/B4 and A5/B5, respectively). CT values and standard deviations (SD) of the liver and lesions were measured, and the corresponding signal-to-noise ratio (SNR) and contrast-to-noise ratio (CNR) were calculated. The edge rise slope (ERS) was determined using ImageJ software by measuring CT values along a line from the liver parenchyma to the lesion. Objective metrics were compared using one-way ANOVA, with independent samples t-tests applied for inter-group differences. Subjective scoring, which encompassed noise level, diagnostic confidence, and lesion margin delineation, was conducted by two radiologists, with differences analyzed using the Kappa test.

**Results:** Objective evaluation revealed a progressive decrease in lesion SD and a progressive increase in SNR and CNR from subgroups A1/B1 to A3/B3. The SD of Group A2 decreased by 57.4% compared to A4, while the SNR and CNR of A2 icreased by 19.3% and 24.6% compared to A4. Although subgroup B2 had a lower SNR than B5, the difference was not statistically significant. SNR and CNR in B2 increased by 24.1% and 11.9%, respectively, compared to B4. ERS gradually decreased from A1/B1 to A3/B3. ERS values in A2 and B2 increased by 27.0% and 39.4%, respectively, relative to A5 and B5. Although A3 had a lower ERS than A1 and A2, all DLIR subgroups exhibited higher ERS than A5; similar trends were observed in Group B. Subjective evaluation indicated good inter-reader agreement (Kappa > 0.61, p < 0.05). As DLIR strength increased, noise scores rose progressively in both groups. However, noise in A2 and B2 was lower than in A4/A5 and B4/B5. Diagnostic confidence and lesion margin delineation scores were highest in A2 and B2, while all subjective scores were lowest in A5 and B5.

**Discussion:** Most prior studies evaluated the liver, vessels, or confirmed that image quality can be guaranteed at low doses. However, there are few studies on specific individual lesions. Therefore, this study aims to investigate specific individual lesions. The details and detection rate were analyzed separately to confirm the clinical acceptability of 2.5-mm DLIR image in different contrast lesions.

**Conclusion:** For both high-and low-contrast hepatic lesions, DLIR provides superior image quality compared to ASIR-V,with the 2.5mm DLIR-M setting being optimal. DLIR-M reduces image noise,improves spatial resolution, and produces images more suitable for diagnostic purposes.

---

Abdomen contrast enhanced computed tomography (CT) has been widely used in clinical practice for a long time now, especially in cancer patients for diagnosis, follow-up and periodic surveillance. The standard slice thickness for abdominal CT is 5 mm[1], which results in images that are relatively low noise and poor detail resolution. Although it is possible to accomplish thinner-slice reconstructions, it greatly increases the image noise. There is a clinical need to develop thinner-slice protocols. These protocols should provide high spatial resolution and good visualization of anatomical details with lower noise. This would enhance detection and characterization of lesions. This is a priority in CT research today[2].

The iterative reconstruction (IR) algorithm is currently the mainstay in clinical applications, with Adaptive Statistical Iterative Reconstruction-V (ASIR-V) being particularly prevalent. However, studies have shown that IR algorithms can introduce a “smooth” or “plastic” texture to images[3-4], and many IR approaches alter both noise magnitude and texture—changes that may adversely affect the diagnosis of low-contrast lesions. In recent years, the Deep Learning Image Reconstruction (DLIR) algorithm has emerged as a promising alternative, attracting growing interest in the field[5]. The DLIR engine utilizes a model based on a deep convolutional neural network (DCNN)[6] to generate output images from the acquired raw projection data. This study employs deep neural networks to replicate the characteristics of high-quality Filtered Back Projection (FBP) images. Consequently, Deep Learning Image Reconstruction (DLIR) retains sharpness and noise texture comparable to FBP[7], while simultaneously enhancing overall image quality[8].Furthermore, DLIR offers three distinct reconstruction strength levels: low, medium, and high. Abdominal space-occupying lesions belong to various types manifesting different enhancement patterns during arterial and portal venous phase which show a varied level of contrast. This research aims to explore the comparative benefits and drawbacks of thin-slice(2.5mm) ASIR-V and DLIR images in CECT of the abdomen for evaluation of high and low-contrast hepatic lesions and the suitable DLIR strength for liver lesion assessment.

## 1. Materials and Methods

### 1.1 Patient Population

A retrospective analysis was conducted on 90 patients who underwent contrast-enhanced abdominal CT at the Second People’s Hospital of Changzhou between February and December 2024. Group A comprised 48 patients with low-contrast lesions (all metastases). Group B comprised 42 patients with high-contrast lesions (40 hemangiomas and 2 cases of focal nodular hyperplasia). The detailed inclusion and exclusion criteria are shown in Fig. 1. This study was approved by the hospital’s Institutional Review Board.

**Fig. 1.**
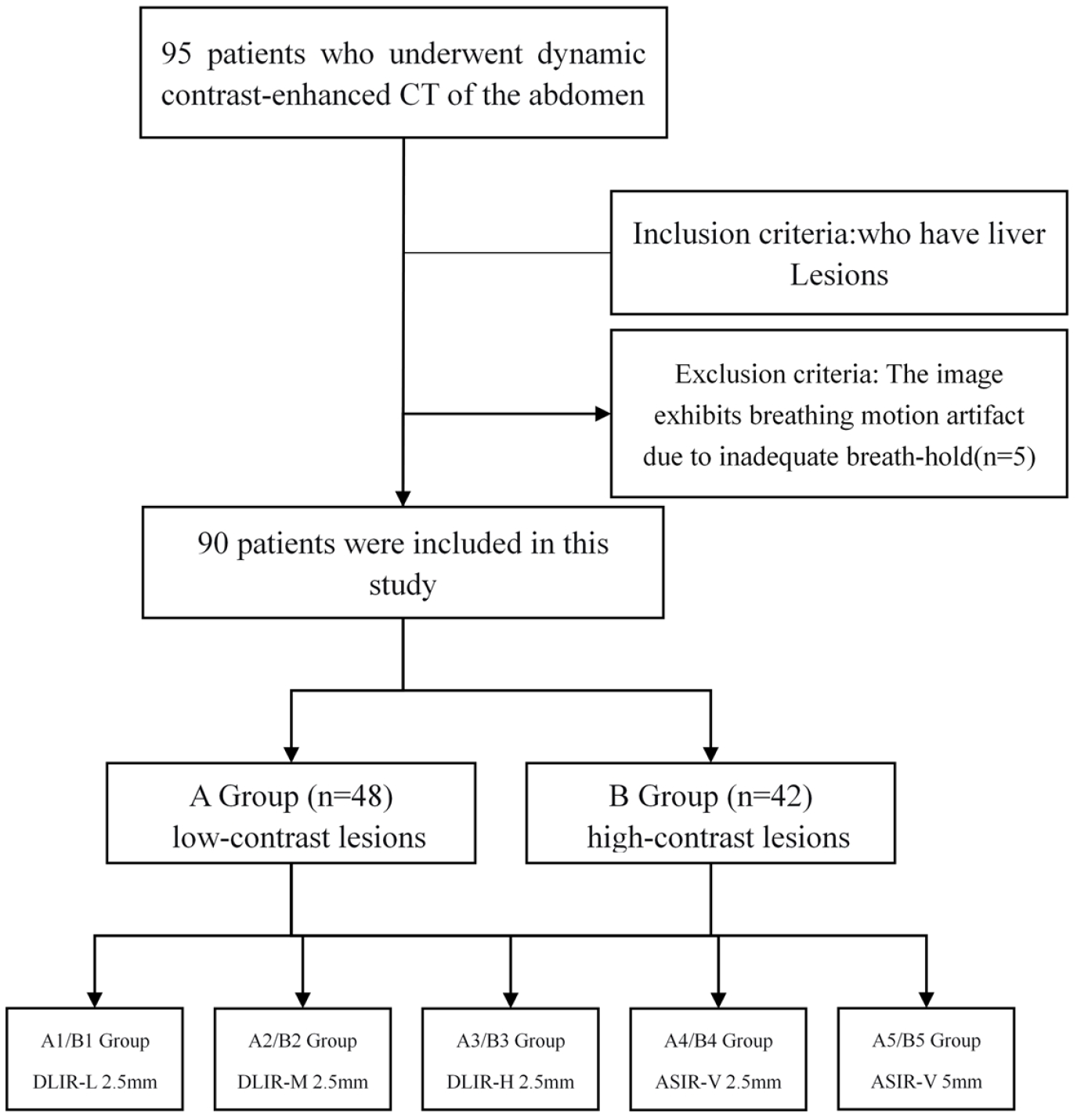
Flowchart of the subject selection and study design

**Fig. 2.**
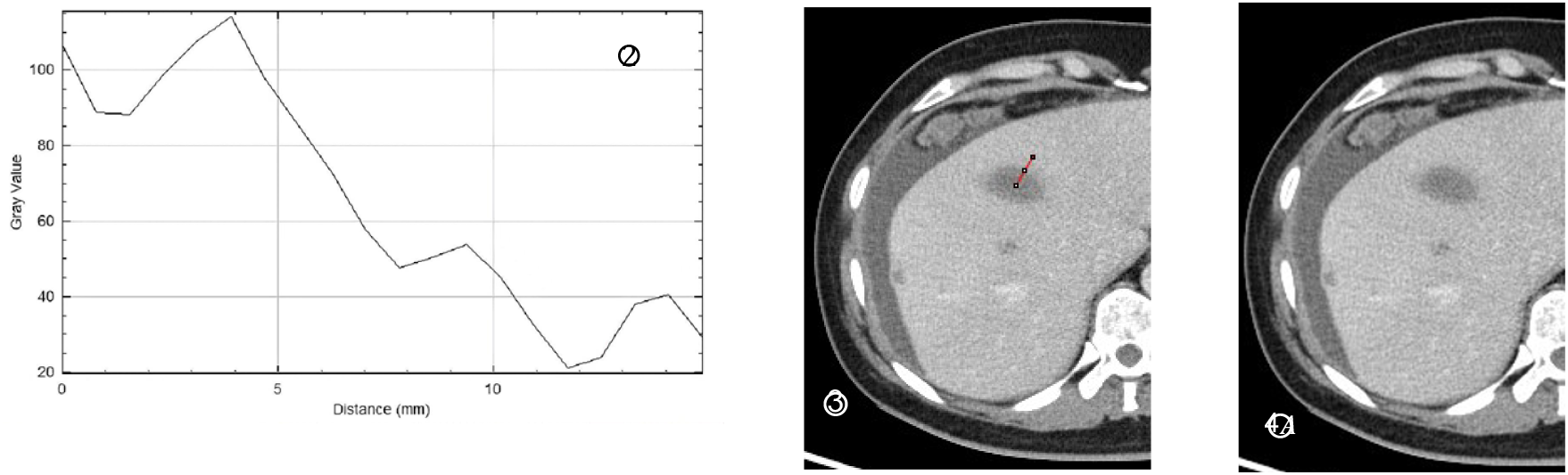
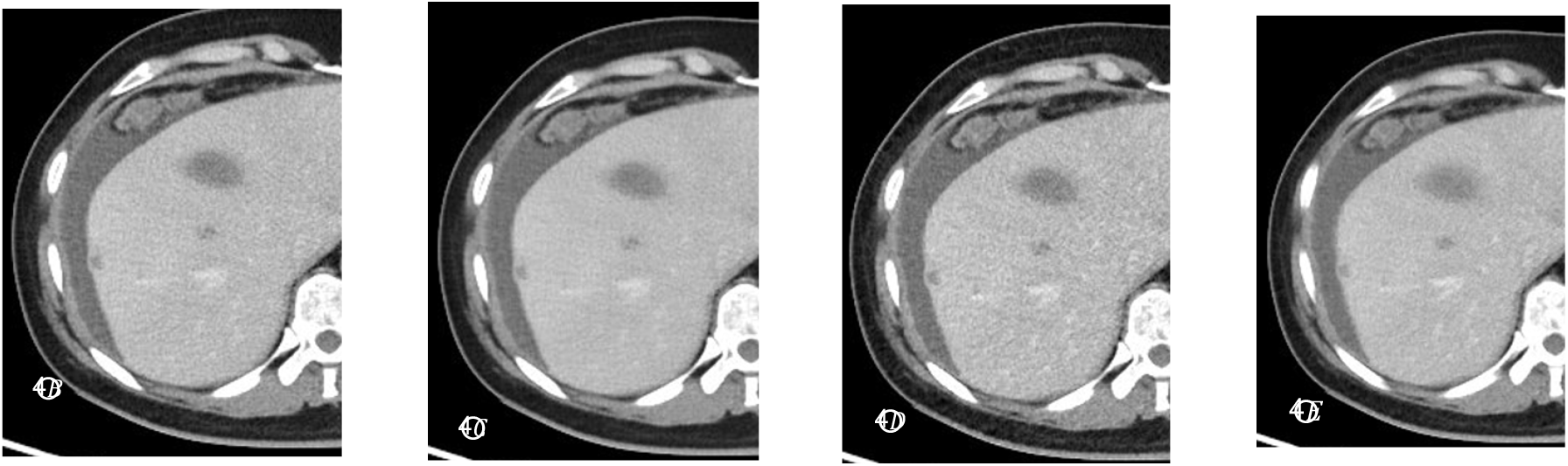
The CT value curves of all points on the line were measured of image J.Fig.3 ImageJ marks the line of measurement Fig.4A∼E The images were reconstruted with 2.5mm DLIR-L, DLIR-M, DLIR-H, 2.5mm ASIR-V 50%, 5mm ASIR-V 50% respectively.

### 1.2 CT Examination Protocol

All examinations were performed using a GE Revolution CT scanner (USA). Scanning was performed in a craniocaudal direction, with the range extending from the diaphragmatic dome to the inferior edge of the liver. A non-ionic contrast agent (Iodixanol, 270 mgI/mL) was administered intravenously via a power injector at a dose of 70 mL, followed by 20 mL saline flush, both at a flow rate of 3 mL/s. Using the SmartPrep technique, bolus tracking was performed by monitoring the abdominal aorta; arterial phase scanning was initiated with a 10-second delay after the trigger threshold of 100 Hounsfield Units (HU) was reached. The portal venous phase scan followed after an additional 33-second delay. Other key scanning parameters were: noise index 10.9, pitch 0.992:1, gantry rotation time 0.5 s/rot, detector coverage 80 mm, and a primary reconstructed slice thickness of 5 mm using 40% ASIR-V.

### 1.3 Image Reconstruction

Portal venous phase images were reconstructed using various post-processing algorithms and parameters. The DLIR was applied at low (DLIR-L), medium (DLIR-M), and high (DLIR-H) strengths, forming subgroups A1/B1, A2/B2, and A3/B3, respectively, all with a uniform slice thickness of 2.5 mm. Additionally, the ASIR-V at 50% was employed to reconstruct images of 2.5 mm and 5 mm slice thicknesses, generating subgroups A4/B4 and A5/B5, respectively.

### 1.4 Image Evaluation

#### 1.4.1 Objective evaluation of Image

For each reconstructed image series, objective analysis was performed on axial images. The CT values and standard deviation (SD) of the lesions and adjacent liver parenchyma were measured. A region of interest (ROI) was placed in the center of the lesion, and care was taken to avoid including surrounding normal tissue. Measurements were performed three times, and the average values were used for calculation.The signal-to-noise ratio (SNR) was calculated as: SNR = CT_lesion_/ SD_lesion_.The contrast-to-noise ratio (CNR) was calculated as: CNR = |CT_lesion_-CT_liver_| / SD_liver_. All data measurements are performed by radiologists with more than ten years of diagnostic experience. All points on the straight line from the liver parenchyma to the lesion were measured using ImageJ software (National Institutes of Health, USA) to mark the measurement points at the same location in each set of images CT value, the ratio of the difference between the trough and the peak on the rapidly rising or falling CT number curve divided by the distance between two points is defined as the edge-rise-slope (ERS), and the larger the ERS value, the higher the spatial resolution of the image(Fig.2 and 3).

#### 1.4.2 Subjective evaluation of Image

Subjective image quality was independently scored by two radiologists, each with more than ten years of experience, who were blinded to the reconstruction techniques. The scoring criteria are detailed in Table 1.

**Tab 1.**
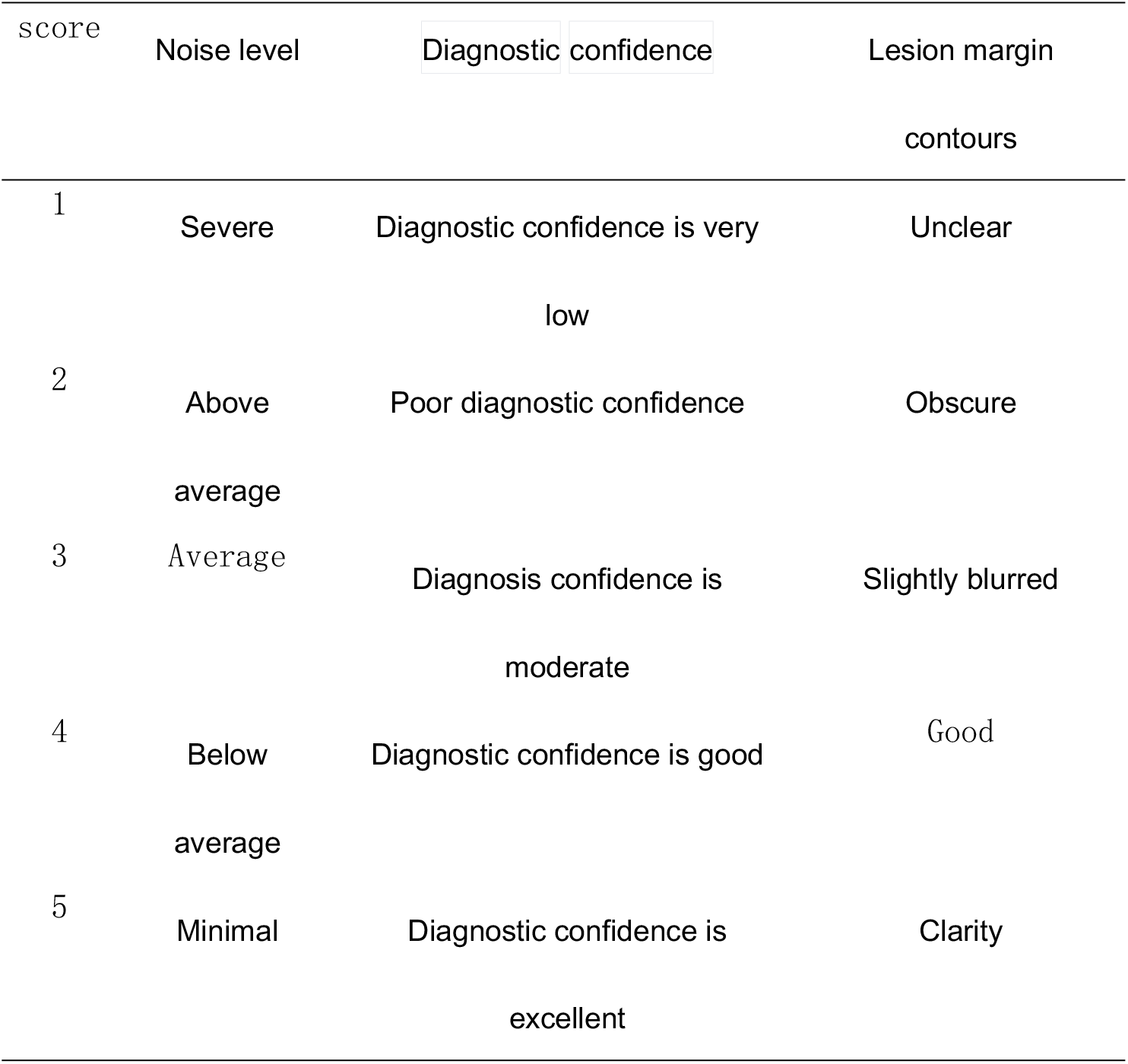
Subjective evaluation of image quality.

#### 1.4.3 Lesion detection evaluation

Two experienced abdominal imagers with over ten years of experience each, set the reference standard for metastatic lesions by reviewing all evidence including extra MRI scans and subsequent CT scans. Subsequently, the lesion detection rates were determined by comparing the lesions on the index images to this reference standard.

### 1.5 Statistical analysis

All data were analyzed with SPSS 24.0, and the data in each group were tested for normality, and the normally distributed data (CT values of lesions, SD, SNR, CNR, and ERS) were expressed in *x*±s and repeated measures ANOVA,and Least Significant Difference was used between groups. The consistency of the subjective scores of the two radiologists was evaluated by the Kappa test, and 0≤ Kappa<0.20 was poor consistency,0.21≤Kappa<0.40 was moderate,0.41≤Kappa<0.61 was moderate,0.61≤Kappa<0.80 was good consistency,and 0.81≤Kappa≤1.0 was very good[9].

## 2 Results

### 2.1 General information

Gender, age, and BMI groups were consistent among group A and B (P>0.5, Table 2).

**Table 2.**
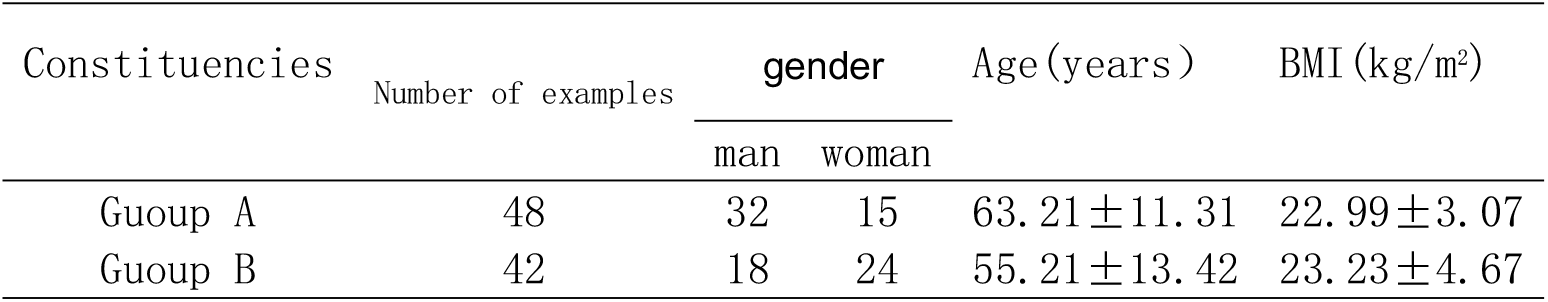
Comparison of the two groups of general data.

### 2.2 Objective evaluation

The objective metrics for group A and B are shown in Tables 3 and 4. There was no difference in lesion CT values between the two groups (p > 0.05). The lesion SD values for groups A4 and B4 were the highest within their respective groups. The lesion SD values for A1–A3 showed a gradually decreasing trend, while the SNR and CNR for A1–A3 gradually increased(Fig.4A∼E). The SD values of groups A2 and A3 were reduced by 19.8% and 57.4% compared with those of group A4, and the SNR and CNR of group A2 were increased by 19.3% and 24.6% compared with group A4.The ERS for groups A1–A3 showed a gradually decreasing trend. The ERS of the group A2 was 27.0% higher than that of the group A5. The ERS of the group A3 was lower than that of the groups A1 and A2, and also lower than the group A4, but the ERS of all these groups (A1-A4) was higher than that of the group A5.The conclusions for the SNR, CNR, SD, and ERS values in groups B1–B3 were the same as those for Group A. The SNR and CNR of the group B2 increased by 24.1% and 11.9%, respectively, compared to the group B4. Although the SNR of the group B2 was lower than that of the group B5, the inter-group comparison showed no statistically significant difference. The ERS of the group B2 was 39.4% higher than that of the group B5. The ERS of group B3 was also lower than that of the group B1 and B2.It is noteworthy that the ERS for groups A3 and B3 was inferior to those of groups A2 and B2, respectively. ?

**Tab 3.**
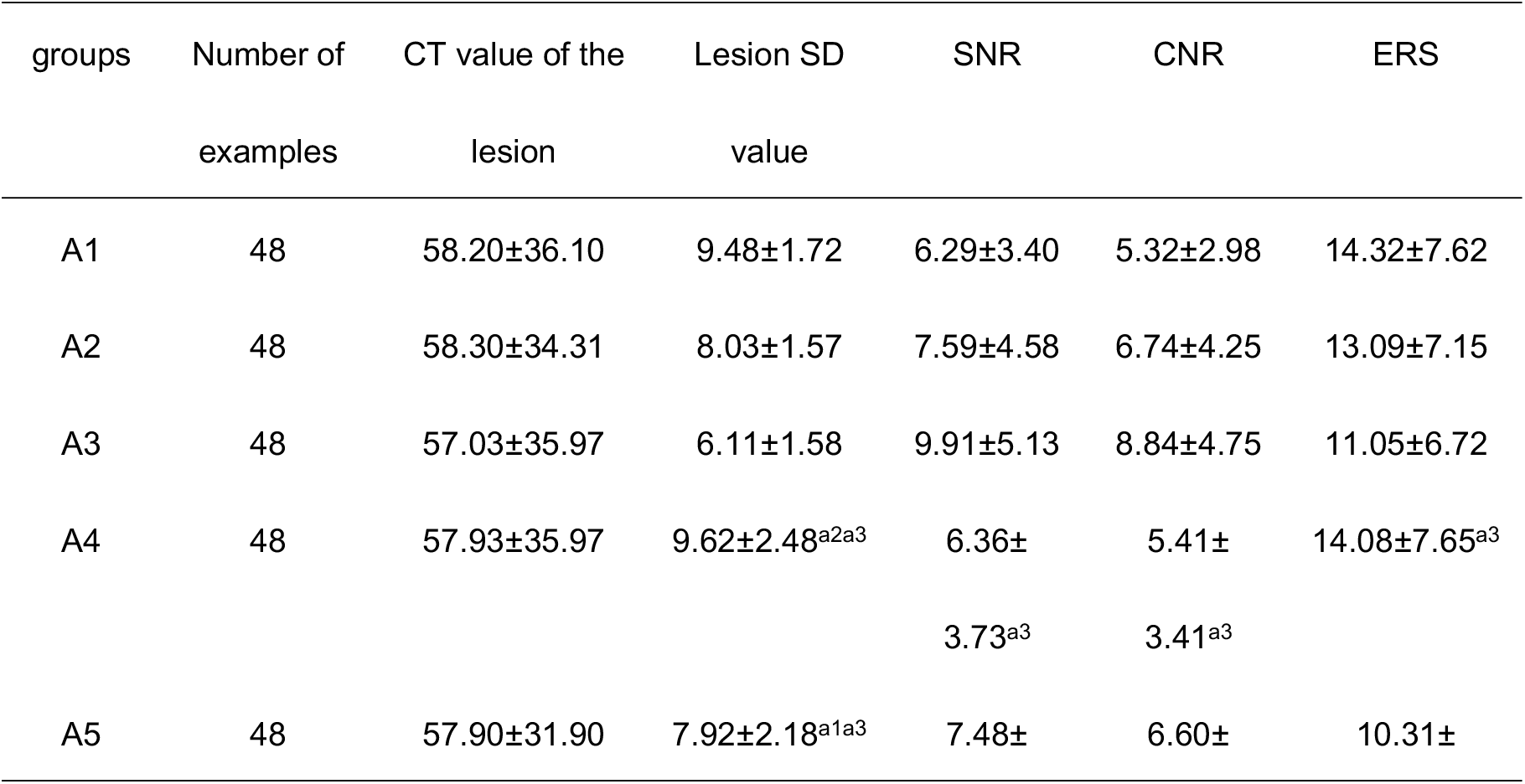
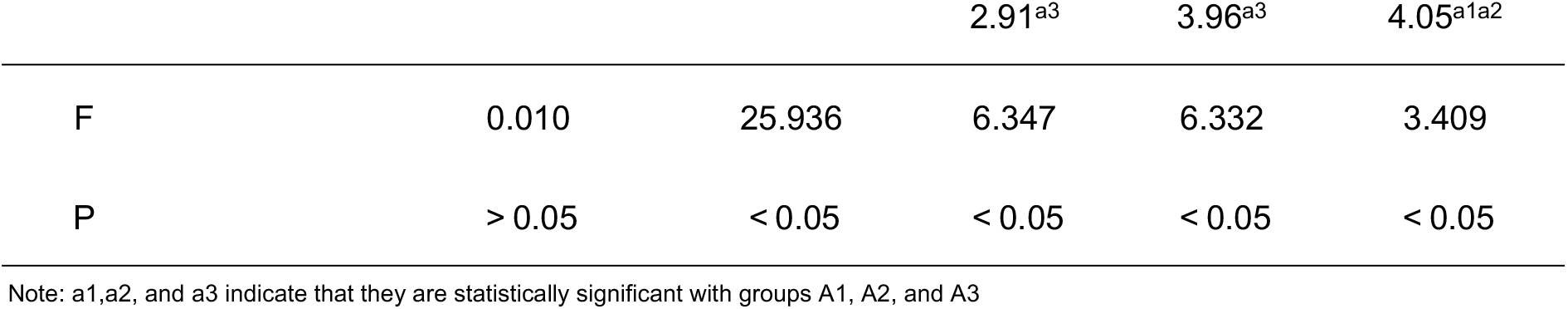
Objective evaluation of the lesions CT values,standard deviation values,SNR,CNR and ERS in different reconstruction methods.

**Tab 4.**
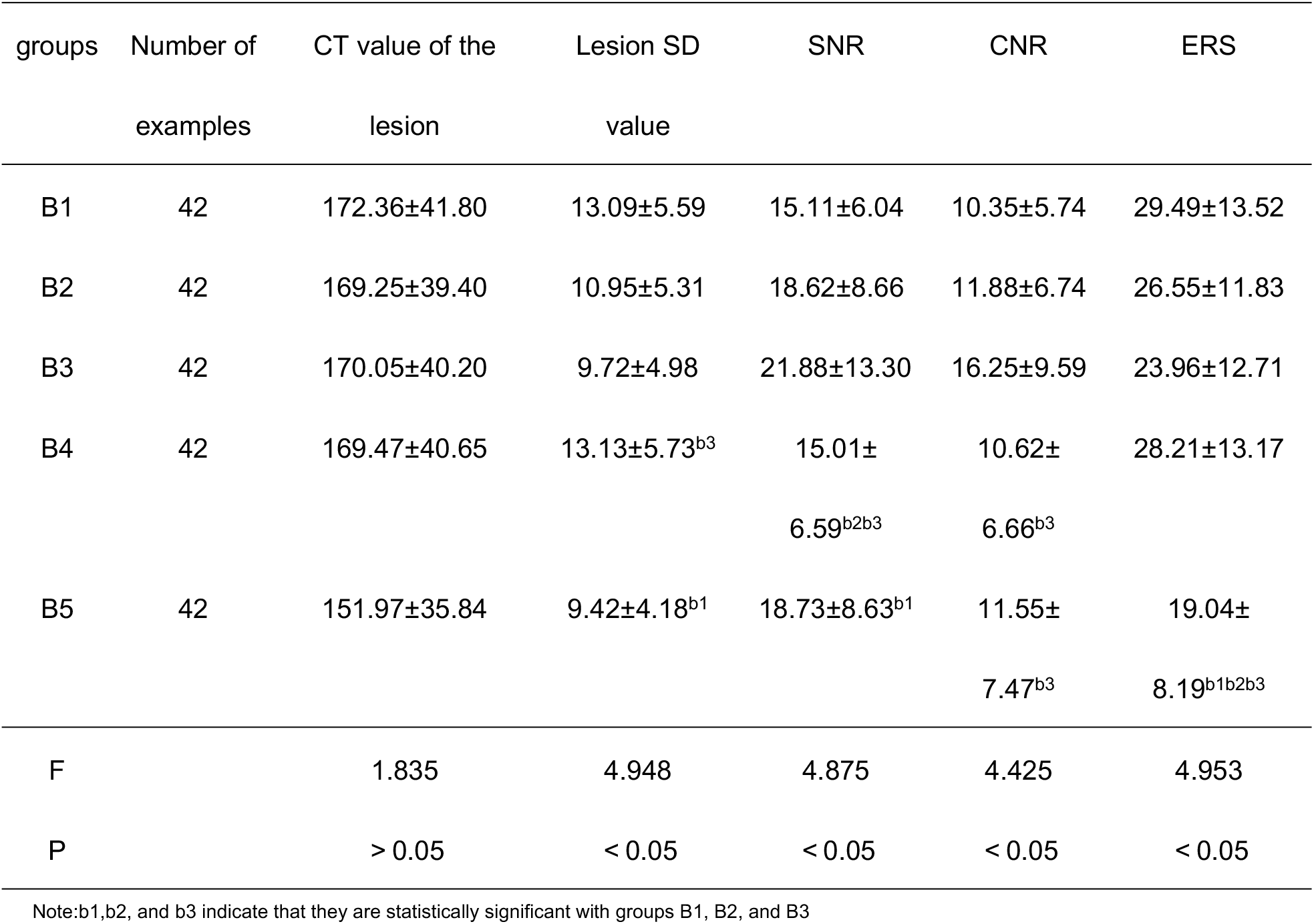
Objective evaluation of the lesions CT values,standard deviation values,SNR,CNR and ERS in different reconstruction methods.

### 2.3 Subjective evaluation

The two physicians had good agreement in subjective scores (Kappa>0.61), which are shown in Tables 5 and 6, and the differences between groups were statistically significant (p<0.05). With increasing DLIR strength, noise scores rose progressively, reaching the highest level in subgroups A3 and B3. Nevertheless, subgroups A2 and B2 demonstrated lower noise than A4/A5 and B4/B5, respectively. Diagnostic confidence and lesion margin delineation scores were highest in subgroups A2 and B2, exceeding those in A3 and B3. The lowest subjective scores across all metrics were observed in subgroups A5 and B5.

**Tab 5.**
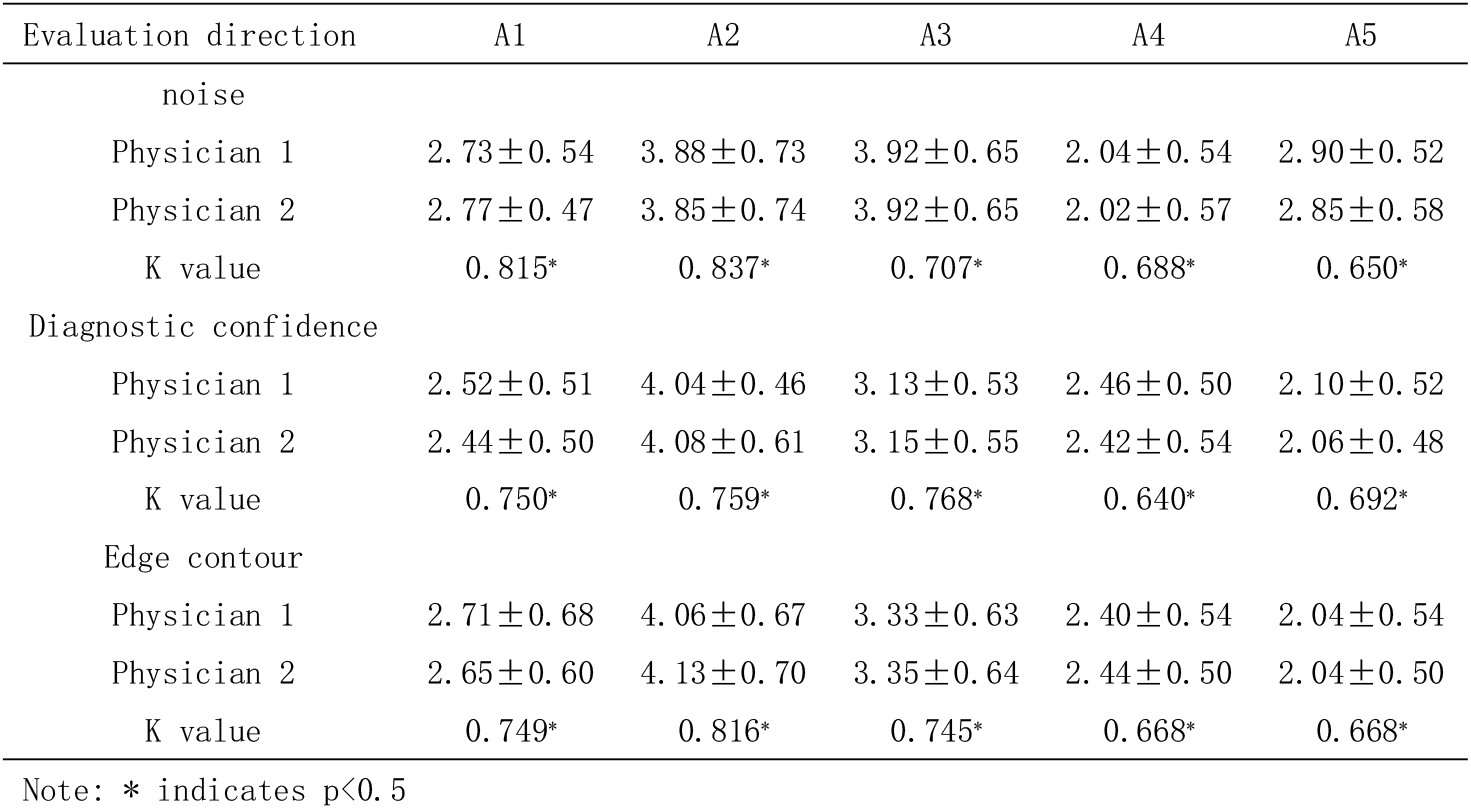
subjective score results of group A.

**Tab 6.**
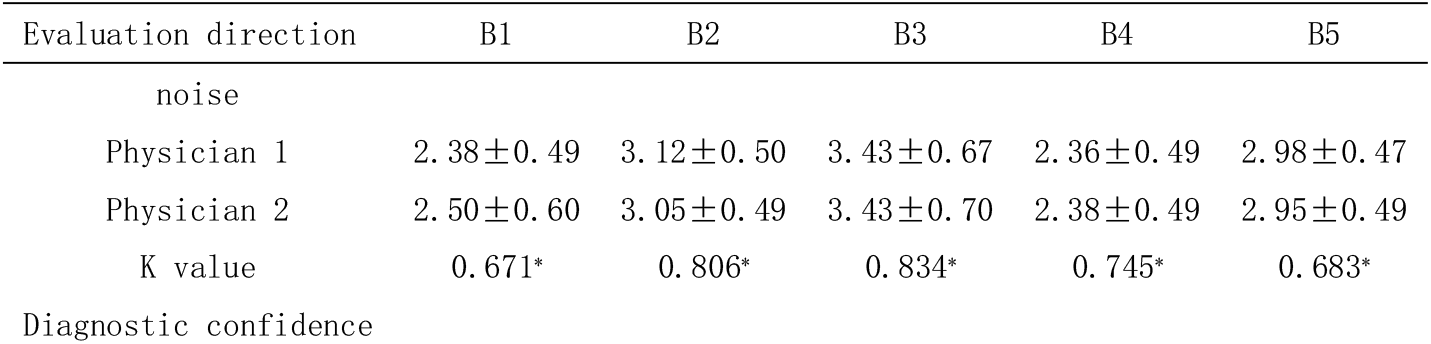
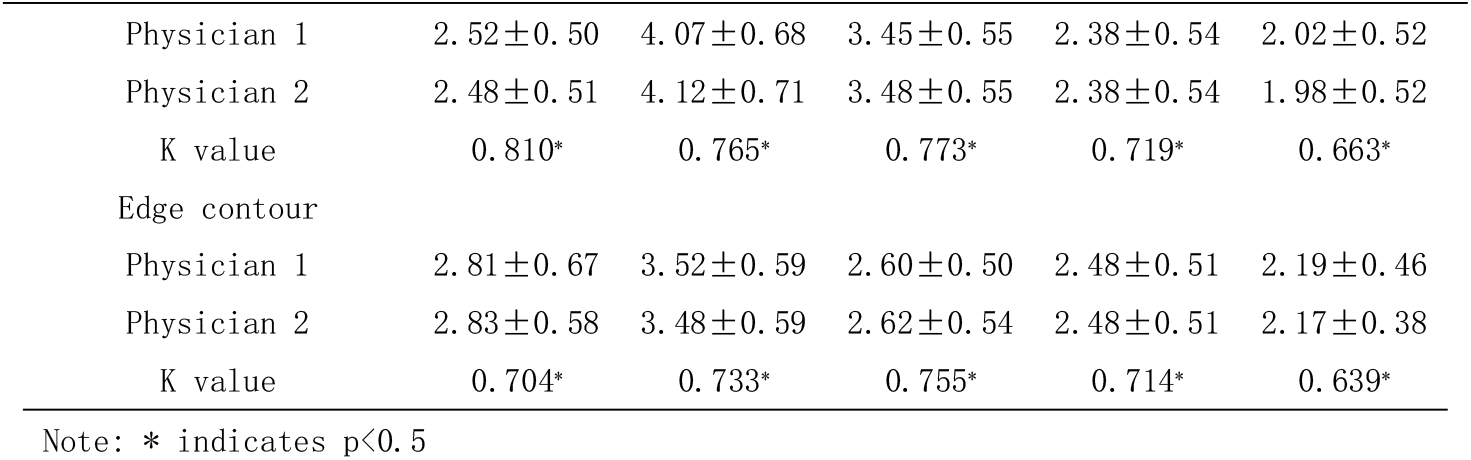
subjective score results of group B.

### 2.4 Lesion Detection Rate

The DLIR groups had higher overall lesion detection rates than the ASIR-V groups for both readers. Reconstruction algorithms did not show any significant difference with sore in lesions larger than 5 mm. In lesions less than 5 mm, the detection rate was highest in A2 and lowest in A5. Upon consensus review, small metastases were diagnosed on lesions missed on initial reading by the 2 radiologists. In Group B (with mostly single lesions), there was a high inter-reader agreement with no difference between the two specialists(Table 7).

**Tab 7.**
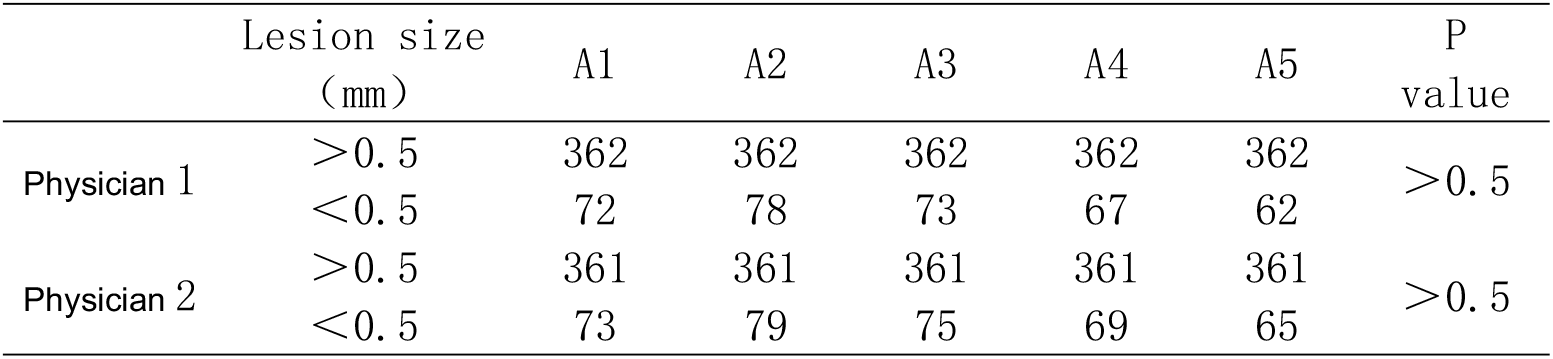
Number of lesions detected by different reconstruction algorithms.

## 3 Discussion

With the development of artificial intelligence (AI), the concept of deep learning was born, and then convolutional neural networks appeared, which improved the image quality without changing the noise texture and solved the problem of image texture change caused by IR reconstruction algorithm [7].

Currently, many studies have confirmed that DLIR can significantly improve the image quality of abdominal CT compared to ASIR-V at low doses [10-[13]. It has been confirmed that 1.25 mm images based on DLIR-M and DLIR-H reconstruction are better than conventional 5 mm ASIR-V reconstructed images, with better image quality and lower image noise [14]. Most prior studies evaluated the liver, vessels, or confirmed that image quality can be guaranteed at low doses. However, there are few studies on specific individual lesions. Therefore, this study aims to investigate specific individual lesions. The details and detection rate were analyzed separately to confirm the clinical acceptability of 2.5-mm DLIR image in different contrast lesions. Most cases studied in this paper were abdominal examination patients. Hence, the resolution of soft tissue was a high one. The 1.25mm image noise being too high, the 2.5mm image was selected for making the comparison.

The results of our investigation demonstrated that as DLIR strength increased, the SD of the lesions progressively decreased, while the SNR and CNR increased in both high- and low-contrast groups, consistent with previous studies[15-16]. Though it was reported in one study that DLIR-H had the best spatial resolution and diagnostic confidence, this would be due to the fact that the evaluation was of liver parenchyma and portal vein only[1]. Conversely, the DLIR may not maintain detection confidence for very small low-contrast lesions[17]. Additionally, one study found that DLIR-M images represented the optimal balance between image quality and diagnostic confidence for lesions in low-dose abdominal CT[18].The DLIR improved hepatocellular carcinoma (HCC) detection without altering noise texture[19], and higher detectability for low-contrast liver lesions with medium- and high-strength DLIR[20]. The metric ERS introduced in this study reflects spatial resolution and shows a decrease in value with increasing levels of DLIR,confirming that the spatial resolution for thin-section images is better than that for thicker sections. The ERS in the group DLIR-M was higher than that in the group DLIR-H for both high- and low-contrast lesions. Although DLIR is designed to reduce noise while preserving texture,our investigation indicates that the highest DLIR strength does not enhance spatial resolution for space-occupying lesions,It is posited that the exceptional noise reduction capability of high-strength DLIR may be achieved at the expense of spatial resolution. The scores for diagnostic confidence and edge sharpness in the subjective evaluation corroborated the findings of the ERS. Furthermore, overall lesion detection was superior in the DLIR groups compared to the ASIR-V groups. While both reconstruction algorithms demonstrated comparable performance for lesions larger than 5 mm, the group DLIR-M achieved the highest detection rate for sub-5 mm lesions-a finding consistent with the previous report[21]. Some studies have cautioned that DLIR may alter intrinsic image properties[22]. In light of this and our own results, high-strength DLIR should be applied cautiously in liver imaging. When considering the balance between noise suppression, spatial resolution, and diagnostic detail, DLIR-M appears to offer the most favorable trade-off. Although subgroups A4 and B4 exhibited a higher ERS than the medium- and high-strength DLIR groups, they performed significantly worse in terms of noise, SNR, and subjective ratings, rendering them unsuitable for routine use. Overall, comprehensive analysis indicates that DLIR yields superior image quality relative to ASIR-V, with DLIR-M being the recommended strength for liver lesion evaluation.

This study has several limitations. First, the potential influence of variations in patient body mass index (BMI) on image quality was not evaluated; future studies should assess this factor. Second, following previous literature indicating its favorable performance in abdominal imaging, ASIR-V 50%[23] was selected as the comparator, while other blending weights of ASIR-V were not explored. Additionally, only a 2.5 mm slice thickness was used for DLIR reconstruction; thinner slices were not compared or further validated.

In summary, regarding liver space-occupying lesions, a comprehensive analysis of both subjective and objective evaluations indicates that the 2.5-mm DLIR-M reconstruction can effectively replace the commonly used ASIR-V algorithm in clinical practice. This replacement results in CT images characterized by reduced noise, enhanced spatial resolution, and improved diagnostic interpretability.

The authors confirm contribution to the paper as follows: Jing Zhang was responsible for the study design, manuscript writing, and review; Li Li performed data collection and organization; Jian Cao conducted the experiment execution; Jie Yang drafted the initial manuscript. All authors read and approved the final manuscript.

## Data Availability

The data in my article can be validation, replication, reanalysis, new analysis, reinterpretation or inclusion into meta-analysesensuring the reproducibility of the study.

